# The nexus of travel restriction, air pollution and COVID-19 infection: Investigation from a megacity of the southern China

**DOI:** 10.1101/2020.04.25.20079335

**Authors:** Wei Li, Xiaohong Chen

## Abstract

To control and prevent the spread of COVID-19, generalized social distancing measures, such as traffic control and travel restriction acted in China. Previous studies indicated that the traffic conditions had significant influence on the air quality, and which was related to the respiratory diseases. This study aimed to reveal the nexus of travel restriction, air pollution and COVID-19. Shenzhen, one of the top 4 megacities in China was considered as the study area, statistical analysis methods, including linear/nonlinear regression and bivariate correlation was conducted to evaluate the relationship of the traffic and passenger population, travel intensity, NO_2_, PM_10_, PM_2.5_ and the number of COVID-19 confirmed cases. The results suggested that traffic control and travel restriction had a significant correlation with the number of COVID-19 confirmed cases, which shown negative correlation with the traffic intensity of the city, NO_2_, PM_10_ and PM_2.5_ show significant positive correlation with the traffic intensity, traffic control and travel restriction would slow down and prevent the spread of the viruses at the outbreak period. Different study scale might results in different results, thus the research focused on the nexus of traffic control and travel restriction, air pollution and COVID-19 should been enhanced in future, and differentiated epidemic control and prevention measures should be considered according to the different situation of cities as well as countries.

## 1. Introduction

Since 31 December 2019 China reported a cluster of cases of pneumonia in Wuhan, Hubei Province (WHO, 2020a; Chen et al., 2020; Xie and Zhu., 2020), a novel coronavirus disease, namely COVID-19 spread rapidly and became a world pandemic (WHO, 2020a,b; Vellingiri et al.. 2020), there have been 2,164,111 confirmed cases of COVID-19 in 213 countries, areas or territories, reported to WHO as of 2:00 am CEST, 18 April 2020 (WHO, 2020b). To prevent and reduce spread of the infection, strong and immediate measures, such as detect disease early, isolate and treat cases, trace contacts and social distancing acted in China at national scale (Pan et al., 2020). Modeling studies indicated that the travel restrict of Wuhan delayed the epidemic progression by 3 to 5 days in mainland China (Chinazzi et al., 2020; Tian et al., 2020; Pan et al., 2020), while reducing case importations to other countries by nearly 80% through mid-February (Chinazzi et al., 2020; Pan et al., 2020).

Previous studies indicated that the respiratory disease was highly related to the air pollution (Phosri et al., 2019; Zhu et al., 2019), and which could be considered as a putative disease risk factor of the novel COVID-19 disease (Martelletti and Martelletti, 2020). Study conducted by Zhu et al., 2020 in China reveled that there was a significant relationship between COVID-19 infection and air pollutants such as PM_10_, PM_2.5_ and NO_2_. To prevent and control the spread of the novel coronavirus disease, social distancing was encouraged in China, people were encouraged to stay at home and limit outdoor activities, the worse epidemic regions took traffic control measures to restrict the travel. Consequently, social distancing was not only a simple polite behavior that keeping distance with people, but also multiple measures, such as traffic control and travel restriction, that controlled and prevented the contact among people and more over cut down the spread of the viruses (Prin and Bartels, 2020; Yezli and Khan, 2020). Different traffic and travel management strategies had a direct effect on air pollution (Pasquier and André, 2017; Krecl et al., 2020), however, to our knowledge, no study has yet comprehensively evaluated the nexus of traffic control and travel restriction, air pollution and COVID-19 infection at city scale, how travel restriction influences human living habit, and the impact on air environment as well as COVID-19 infection should be well concerned.

In this paper, a contradistinctive study was conducted among the changing of intracity and intercity travel restriction, air environment quality and COVID-19 infection in Shenzhen, one of the top 4 megacity of China, from January to March in 2019 and 2020, and the relationship of which was evaluated using statistical methods. The authors hope the findings will provide useful enlightenment for the control and prevention of this novel coronavirus disease.

## 2. Data acquisition and methods

This study was conducted in Shenzhen, southern China, with longitude from 113°43'E to 114°38'E, latitude from 22°24'N to 22°52'N. Shenzhen is one of the top 4 megacities of China, with 1997.47 km^2^ urban area and 13.03 million permanent resident populations. In 2018 the total GDP of Shenzhen was 2422.2 billion CNY and the economic aggregate ranked the top 5 of the Asian cities.

The air quality data were adapted from the national air quality publishing platform (http://106.37.208.233:20035/), hourly data of NO_2_ (ug/m^3^), PM_10_ (ug/m^3^) and PM_2.5_ (ug/m^3^), from 11 stations were collected during 1 January and 31 March in 2019 and 2020. The traffic data, including intracity traffic (the monthly passenger population, 10^4^ person/time) and intercity traffic (the monthly public transport population, 10^4^ person/time) were adapted from the Transport Bureau of Shenzhen Municipality (http://jtys.sz.gov.cn/zwgk/sjfb/index.htm). The travel intensity (TI) of Shenzhen was collected from Baidu Qianxi (http://qianxi.baidu.com/), an open data platform based on the location based service (LBS) big data technology, and the daily TI was the logarithmic results of the ratio of the travel population and the resistant population (cited from the data declaration of the Baidu Qianxi website). The data of COVID-19 confirmed cases were collated from the Shenzhen Municipal Health Commission (http://wjw.sz.gov.cn/yqxx/202004/t20200420_19176831.htm).

The descriptive statistical analysis of the intercity and intracity traffic and travel, air pollutants and COVID-19 confirmed cases was conducted firstly and nonparametric test (Kolmogorov-Smirnov method) (Zhao et al., 2017) was used to reveal the data distribution characters. Secondly, linear/nonlinear regression and bivariate correlation analysis (Grange et al., 2016) was conducted to evaluate the relationship of these variables, and the correlation was considered significant at the 0.01 level (2-tailed) (Zhu et al., 2020).

## 3. Results

### 3.1 data descriptive

Fig. 1 illustrated the temporal variability of the number of COVID-19 confirmed cases and the travel intensity of Shenzhen in 2019 and 2020. With the developments on the novel coronavirus disease in Shenzhen, the time series of January to March 2020 were divided into 3 parts: the normal period, the social distancing period and the reopened period. The normal period was from 1 January to 23 January. During this period, the variation tendency of travel intensity in 2020 was similar with that in 2019. The average value of travel intensity in 2020 was 4.34, with 25% increased from 2019, indicating a more active travel desire of the population in 2020. The social distancing period was from 24 January to 9 February. With the rapidly increasing confirmed cases, Shenzhen strengthened social distancing and the control of traffic and travel, for instance, people were encouraged to stay at home, real-name registration with residential address and personal health condition was conducted by mobile phone APP, and the electric health QR code was checked as the certificate for entering a housing estate (Fig. 2 (a)), some bus routes were closed, the subway and taxi adjusted the serving time and encouraged the passengers to scan the code to record the travel information (Fig. 2 (b) and (c)). During the social distancing period, a sharp decrease of travel intensity appeared, the average value of which was 0.87 (2.21 in 2019 at the same time), decreasing nearly 80% from the normal period. The reopened period was from 10 February to 31 March, the economy and travel was reopened, the travel intensity was slightly increasing with average value of 2.87 (2.3 times of that in the social distancing period), but which was only about 59% of the value in the same time period in 2019, indicating people’s travel desire was not recovered yet.

**Fig. 1.**
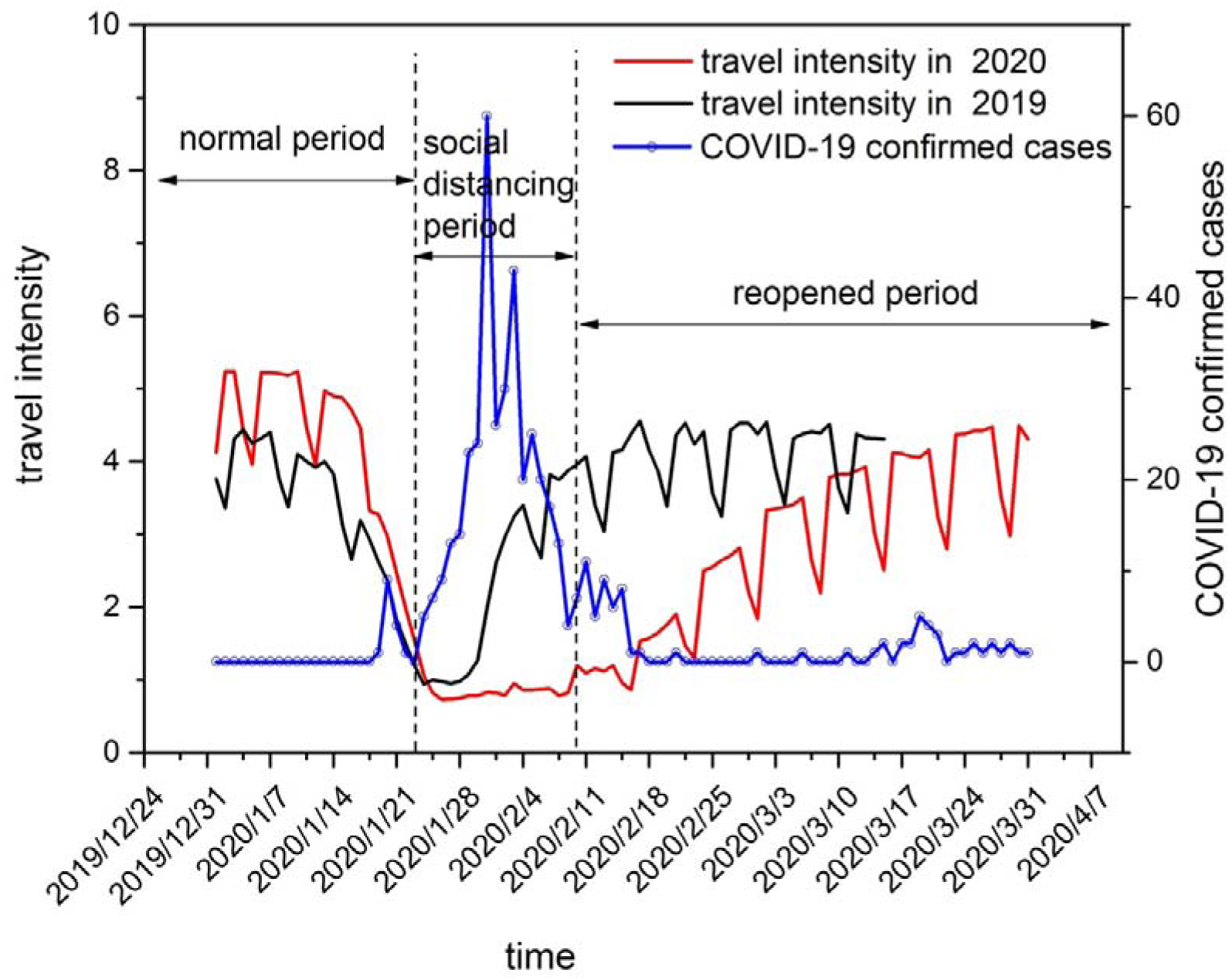
Temporal variability of the confirmed cases and travel intensity

**Fig. 2.**
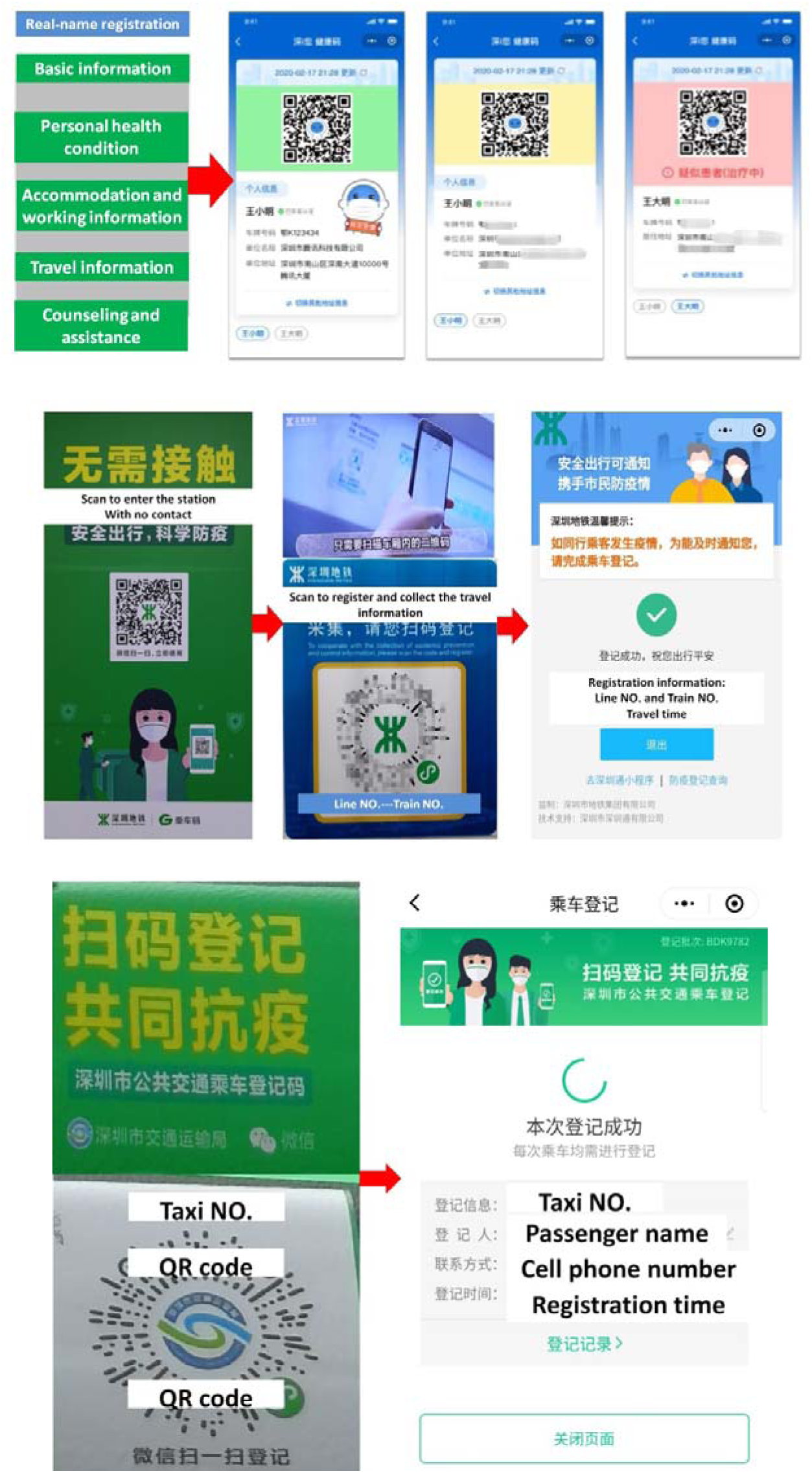
Social distancing, traffic control and travel restriction measures conducted in Shenzhen (a) real-name registration and information collection (b) registration and information collection on the subway (c) registration and information collection on the texi

The intracity traffic (the monthly passenger population) and intercity traffic (the monthly public transport population) revealed the similar situation of the traffic and travel conditions of Shenzhen. In January 2020, the total volume of passenger population (including passengers transferred by civil aviation, highway, rail way and waterway) was about 17.73 million person/times, decreasing about 6.04% compared to 2019, while which was 3.75 million person/times in February 2020, decreasing about 78.82% and 39.48% compared to January in 2020 and February in 2019, respectively. In March 2020, the total volume of passenger population was 6.79 million person/times, increasing about 80.66% compared to February in 2020, but which decreased about 45.89% compared to March in 2019. The total volume of public transport population (including intercity passengers transferred by bus, subway taxi and tramcars) changed rapidly in the same period. In January 2020 the total volume was about 250 million person/times, while it was 41.77 million person/times in February 2020, decreasing about 83.29% and 48.23% compared to January 2020 and February 2019, respectively.

**Fig. 3.**
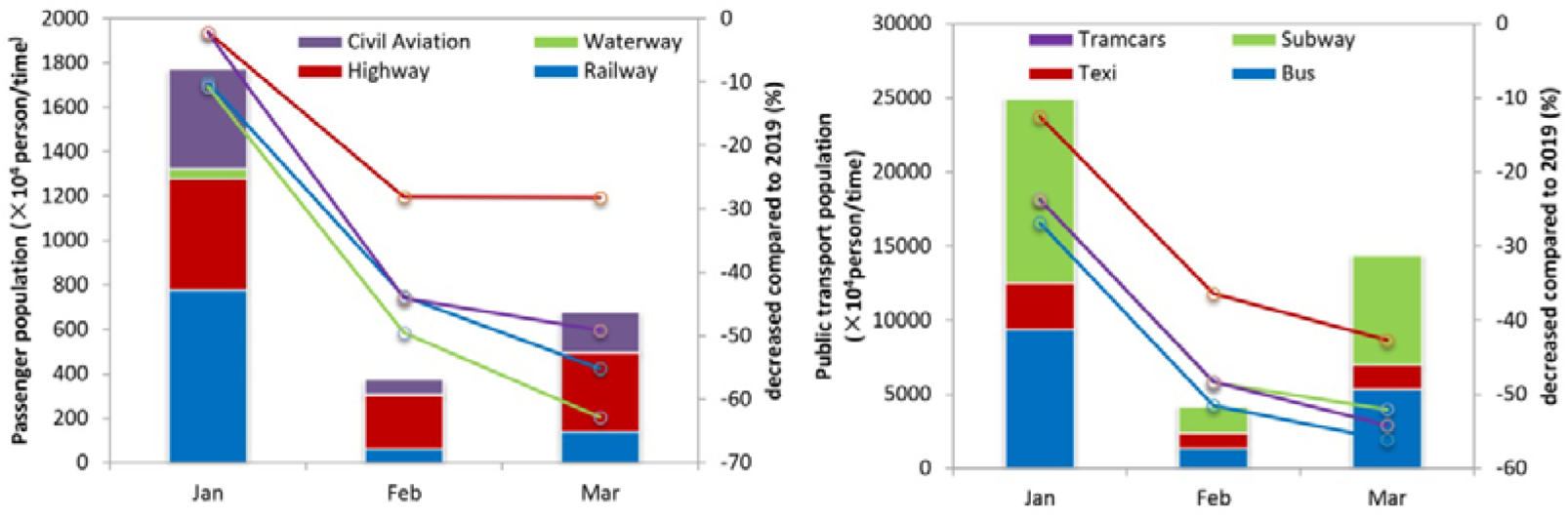
The volume and variable ratio of monthly passenger population and public transport population in Shenzhen from January to March in 2020

With rapidly decreasing of the volume of transport population, the air quality of Shenzhen improved in 2020. Fig. 4 illustrated the concentration variability of NO_2_, PM_10_ and PM_2.5_, both of which decreased compared to 2019. The mean concentrations of NO_2_, PM_10_ and PM_2.5_ in 2020 from January to March were 21.82, 37.54 and 22.53 ug/m^3^, while which were 28.51, 44.52 and 28.02 ug/m^3^, respectively (Fig. 4 (a)). More specifically, the concentrations of air pollutants varied in the similar tendency of the volume of transport population in different time period of 2020. In normal period, the concentrations of NO_2_, PM_10_ and PM_2.5_ were relatively higher with mean value of 28.77, 52.77 and 30.86 ug/m^3^, while in social distancing period the mean value of which were 12.39, 26.83 and 19.33 ug/m^3^ (Fig. 4 (b)), with an average decreasing ratio about 50%. In reopened period, the concentrations of NO_2_, PM_10_ and PM_2.5_ slightly increased with mean value of 22.16, 34.75 and 20.06 ug/m^3^, and which were both lower than the monitored results of the normal period in 2020 and the same time period in 2019.

**Fig. 4.**
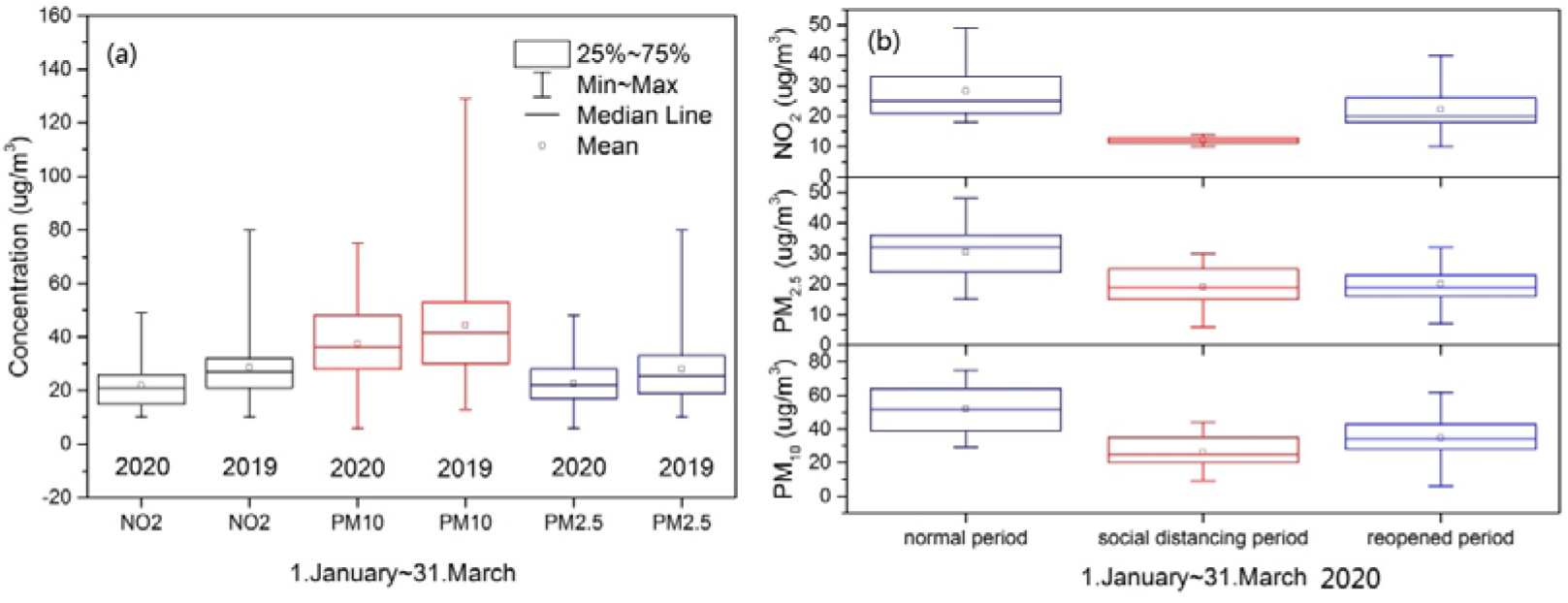
The concentration variability of NO_2_, PM_10_ and PM_2.5_ ((a) the comparative results in 2020 and 2019; (b) the concentration of the air pollutants in different time period in 2020)

### 3.2 correlation and regression analysis of traffic control and travel restriction, air pollution and COVID-19 infection

To reveal the relationship of traffic control and travel restriction, air pollution and COVID-19 infection, both regression and correlation analysis among the travel intensity, concentration of NO_2_, PM_10_, PM_2.5_ and the number of confirmed cased were conducted. The one sample Kolmogorov-Smirnov method test was used to evaluate the variable data distribution. The results indicated that the distribution of NO_2_, PM_10_, PM_2.5_ and travel intensity was normal with standard deviation of 8.5, 14.85, 8.58 and 1.52, respectively, while the distribution of the number of COVID-19 confirmed cases was not normal. Thus, Spearman’s correlation test, a wildly used correlation method was conducted to analyze the relationship of the variables (Wang et al., 2019; Ma et al., 2020). The results indicated that the number of COVID-19 confirmed cases shown significant negative correlation with travel intensity (Table 1), with correlation coefficient -0.549 (0.01 level, 2 tailed), indicating reverse variable tendency of travel intensity and the number of COVID-19 confirmed cases, which mean that traffic control and travel restriction would slow down and prevent the spread of the viruses at the outbreak period. The concentration of NO_2_ also shown a significant negative correlation with the number of COVID-19 confirmed cases (correlation coefficient -0.433, 0.01 level, 2 tailed), a possible reason was that the increasing number of COVID-19 confirmed cases inhibited the travel desire, and decreased volume of traffic resulting in the reduction of NO_2_ emissions. The travel intensity shown significant positive correlation with the concentrations of NO_2_, PM_10_, and PM_2.5_, and the correlation coefficients were 0.765, 0.569 and 0.354 (0.01 level, 2 tailed), respectively, suggesting traffic-related air pollution in Shenzhen. The air pollutants also had shown significant positive correlation with each other. The correlation coefficient between NO_2_ and PM_10_, NO_2_ and PM_2.5_ were 0.556 and 0.411 (0.01 level, 2 tailed), while the correlation coefficient between PM_10_ and PM_2.5_ was 0.906 (0.01 level, 2 tailed).

**Table 1.** Spearman’s correlation test results of the variables

|  |  |  | travel<br>intensity | confirmed<br>cases | NO2 | PM10 | PM2.5 |
| --- | --- | --- | --- | --- | --- | --- | --- |
| Spearman's<br>rho | travel<br>intensity | Correlation<br>Coefficient | 1.000 | -.549** | .765** | .569** | .354** |
|  |  | Sig.<br>(2-tailed) |  | .000 | .000 | .000 | .001 |
|  |  | N | 91 | 73 | 91 | 91 | 91 |
|  | confirmed<br>cases | Correlation<br>Coefficient | -.549** | 1.000 | -.433** | -.115 | .090 |
|  |  | Sig.<br>(2-tailed) | .000 |  | .000 | .332 | .451 |
|  |  | N | 73 | 73 | 73 | 73 | 73 |
|  | NO2 | Correlation<br>Coefficient | .765** | -.433** | 1.000 | .556** | .411** |
|  |  | Sig.<br>(2-tailed) | .000 | .000 |  | .000 | .000 |
|  |  | N | 91 | 73 | 91 | 91 | 91 |
|  | PM10 | Correlation<br>Coefficient | .569** | -.115 | .556** | 1.000 | .906** |
|  |  | Sig.<br>(2-tailed) | .000 | .332 | .000 |  | .000 |
|  |  | N | 91 | 73 | 91 | 91 | 91 |
|  | PM2.5 | Correlation<br>Coefficient | .354** | .090 | .411** | .906** | 1.000 |
|  |  | Sig.<br>(2-tailed) | .001 | .451 | .000 | .000 |  |
|  |  | N | 91 | 73 | 91 | 91 | 91 |

\*\* . Correlation is significant at the 0.01 level (2-tailed).
\* . Correlation is significant at the 0.05 level (2-tailed).

The nonlinear regression of travel intensity and COVID-19 confirmed cased was illustrated in Fig. 6, the results fitted with cubic equation and the R^2^ equaled to 0.502 (n=73). The travel intensity and the air pollutants fitted with linear regression, and the slopes were positive while R^2^ equaled to 0.34 (travel intensity with PM_10_), 0.52 (travel intensity with NO_2_) and 0.14 (travel intensity with PM_2.5_), indicating a relatively strong correlation, and these results were in accordance with the Spearman’s test results. The number of COVID-19 confirmed cases and the air pollutants fitted with nonlinear regression, with R^2^ equaled to 0.25 (confirmed cases with NO_2_), 0.01 (confirmed cases with PM_10_) and 0.07 (PM_2.5_), indicating a relatively weak correlation.

**Fig. 5.**
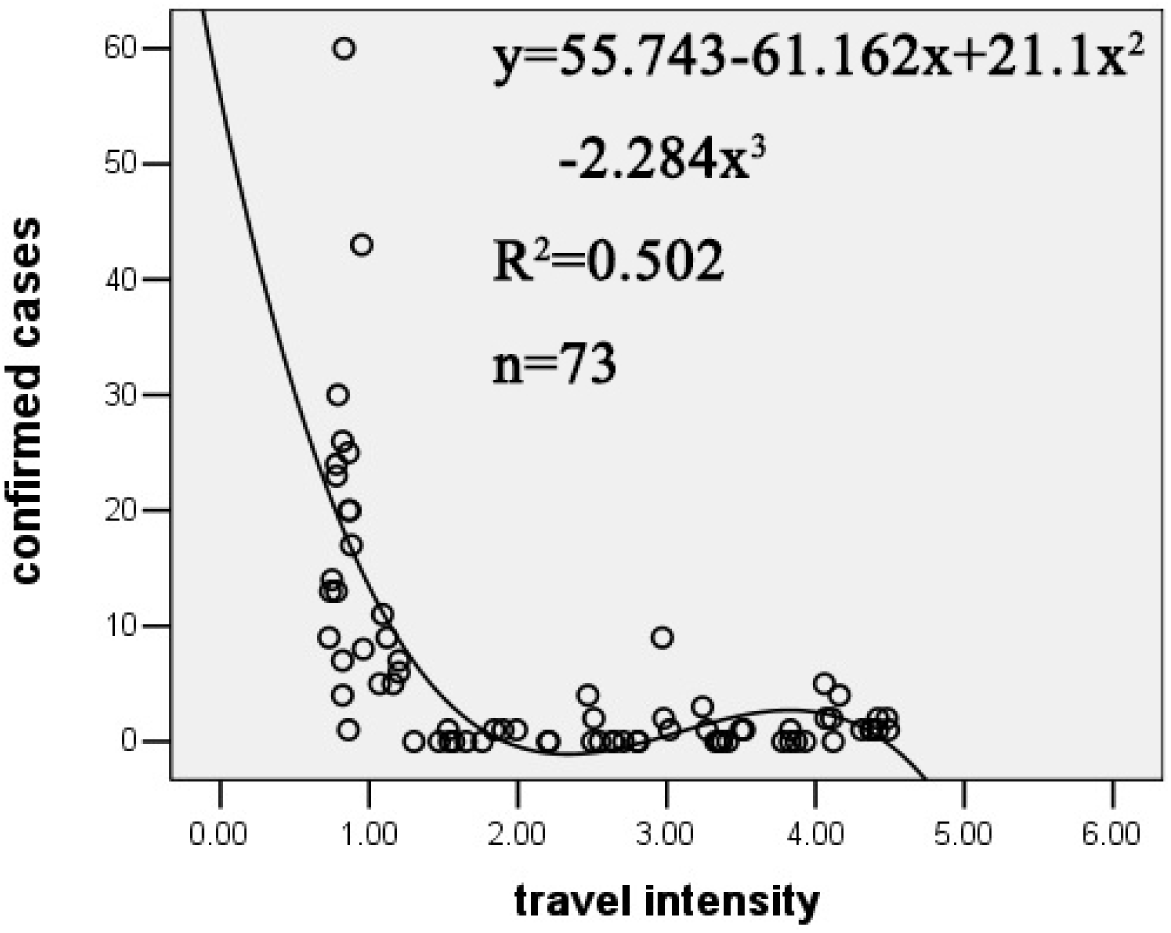
Cubic regression result of COVID-19 confirmed cased and travel intensity

**Fig. 6.**
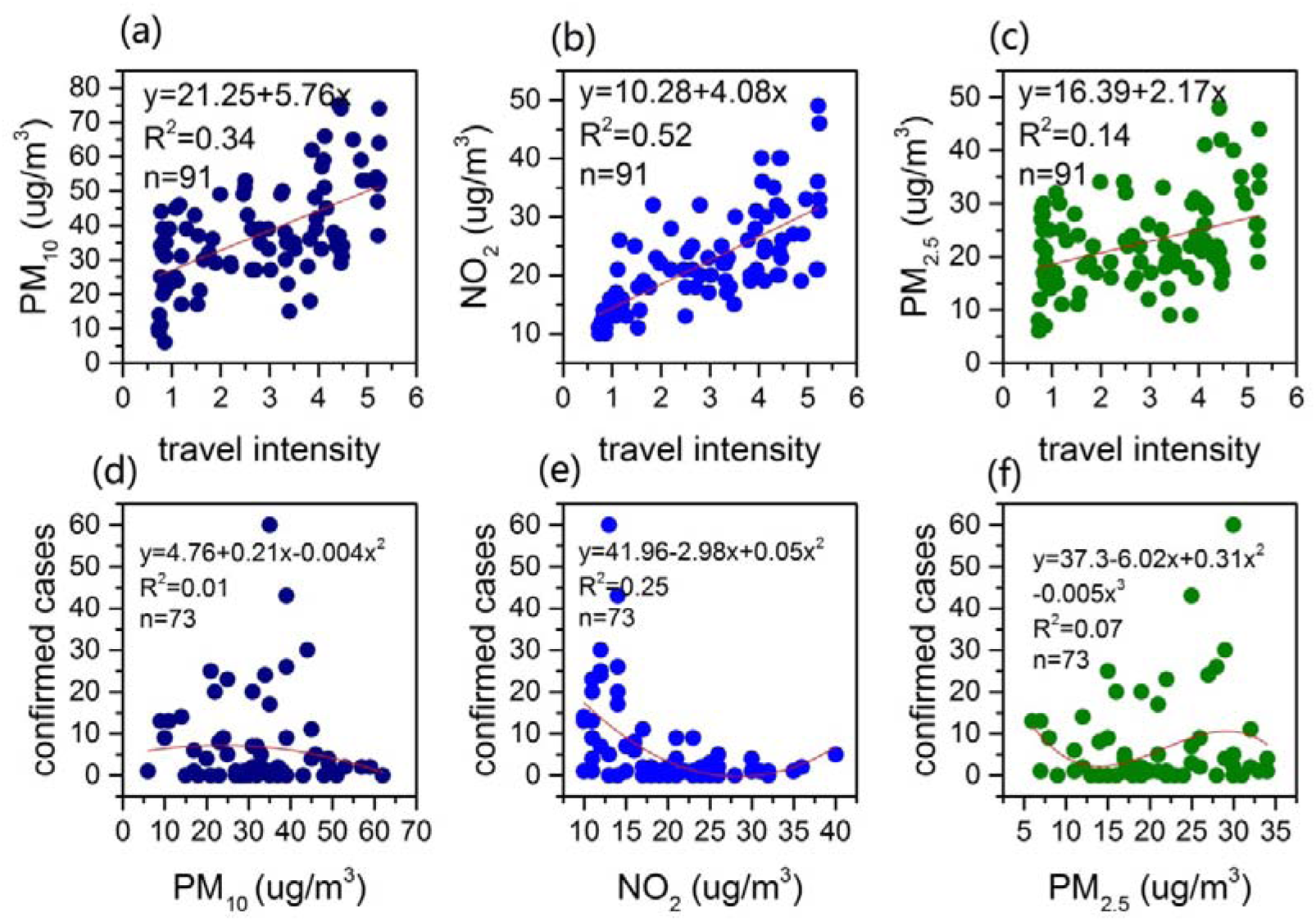
Regression result of air pollutants, COVID-19 confirmed cased and travel intensity

## 4. Discussion and perspectives

The high agglomeration of air pollutants could facilitate the spread of the virus (Martelletti and Martelletti, 2020), the study conducted by Cui et al., 2006 indicated that the mortality rate of SARS shown linear relationship with the API (Air Pollution Index), increasing PM_10_, PM_2.5_, NO_2_, and O_3_ shown associated increasing number of daily COVID-19 confirmed cases in 120 cities of China (Zhu et al., 2020), and the results in this study indicated a significant correlation of the number of COVID-19 confirmed cases and the traffic intensity of Shenzhen, and which had positive correlation with the air pollutants. Thus, the generalized social distancing measures, such as traffic control and travel restriction, although being considered as bold measures facing of political, economic, social and religious challenges (Yezli and Khan, 2020), could be an efficient way to control and prevent the spread of the viruses as well as improving the air quality. On the other hand, the air pollution conditions could be a possible indicator for people to plan the travel during the epidemic prevention and control period.

Besides, previous study conducted by Zhu et al. in the 120 cities of China indicated that air pollutants shown significant correlation with COVID-19 confirmed cases, however, the relationship of the confirmed cases with PM_10_ and PM_2.5_ was weak in this study. Possible reasons to explain were: first, Shenzhen was one of the best air quality cities in China, the relatively low concentrations and little fluctuation of the air pollutants in Shenzhen weakened the relationship of the variables statistically; second, there might be a “scale effect” on the research results, which mean the different scale of the study area and amount of data might result in different results; third, most of the studies focused on the relationship between air pollution and respiratory diseases indicated that the health risk was positive correlation with the exposure concentration of air pollutants and the exposure time (Krecl et al., 2020; Phosri et al., 2020), the while the infection time of COVID-19 was relatively short (news reported that a man with no mask was infected within 15 seconds while shopping in a food market, https://www.thepaper.cn/newsDetail_forward_5844448. ), thus the correlation of air pollutants and COVID-19 infection should be evaluated more accurately considering the COVID-19 infection and pathogenesis as well as the air quality conditions. In order to reveal this, further studies focused on the nexus could be considered to follow two routes: (1) the nexus of these variables among different cities and countries, especially the cities with wildly different air quality conditions, (2) a comprehensive study in one city, regarding the differences of the source of the air pollutants, different travel restriction and traffic control measures and COVID-19 infection and pathogenesis. Moreover, differentiated epidemic control and prevention measures should be considered according to the different situation of cities as well as countries.

Our study had several limitations. First, the travel intensity data was processed daily data which could not reveal the variability of traffic and travel population more accurately, hourly or minutely traffic volume data of the main artery of the city were more useful for further analysis. Second, this study was conducted with statistical method, the meteorological conditions were not considered. Third, this study was conducted at city scale owing to the lack of sufficient data of other cities and countries. Further studies should overcome these limitations and challenges, and the authors hope this study could bring the researcher’s attention on the nexus of the generalized social distancing, air pollution and COVID-19 infection, and conquer this novel disease.

## 5. Conclusion

Generalized social distancing, including traffic control and travel restriction has significant correlation with the number of COVID-19 confirmed cases, which shows negative correlation with the traffic intensity of the city, thus traffic control and travel restriction would slow down and prevent the spread of the viruses at the outbreak period. NO_2_, PM_10_ and PM_2.5_ show significant positive correlation with the traffic intensity, the research focused on the nexus of social distancing, air pollution and COVID-19 should been enhanced in future.

## Data Availability

all data referred to in the manuscript are available on line. The air quality data is collected from https://www.zq12369.com/environment.php. The traffic data, including intracity traffic (the monthly passenger population, 104 person/time) and intercity traffic (the monthly public transport population, 104 person/time) were adapted from the Transport Bureau of Shenzhen Municipality (http://jtys.sz.gov.cn/zwgk/sjfb/index.htm). The travel intensity (TI) of Shenzhen was collected from Baidu Qianxi (http://qianxi.baidu.com/).
The data of COVID-19 confirmed cases were collated from the Shenzhen Municipal Health Commission (http://wjw.sz.gov.cn/yqxx/202004/t20200420_19176831.htm).

http://jtys.sz.gov.cn/zwgk/sjfb/index.htm

http://qianxi.baidu.com/

http://wjw.sz.gov.cn/yqxx/202004/t20200420_19176831.htm

## Acknowledgements

The authors are grateful to Baidu qianxi (http://qianxi.baidu.com/) that provides the city travel intensity data for free.

